# Multi-omics Characterization of Epigenetic and Genetic Risk of Alzheimer Disease in Autopsied Brains from two Ethnic Groups

**DOI:** 10.1101/2024.02.12.24302533

**Authors:** Yiyi Ma, Dolly Reyes-Dumeyer, Angel Piriz, Patricia Recio, Diones Rivera Mejia, Martin Medrano, Rafael A. Lantigua, Jean Paul G. Vonsattel, Giuseppe Tosto, Andrew F. Teich, Benjamin Ciener, Sandra Leskinen, Sharanya Sivakumar, Michael DeTure, Duara Ranjan, Dennis Dickson, Melissa Murray, Edward Lee, David A. Wolk, Lee-Way Jin, Brittany N. Dugger, Annie Hiniker, Robert A. Rissman, Richard Mayeux, Badri N. Vardarajan

## Abstract

**Background:** Both genetic variants and epigenetic features contribute to the risk of Alzheimer’s disease (AD). We studied the AD association of CpG-related single nucleotide polymorphisms (CGS), which act as the hub of both the genetic and epigenetic effects, in Hispanics decedents and generalized the findings to Non-Hispanic Whites (NHW) decedents.

**Methods:** First, we derived the dosage of the CpG site-creating allele of multiple CGSes in each 1 KB window across the genome and we conducted a sliding window association test with clinical diagnosis of AD in 7,155 Hispanics (3,194 cases and 3,961 controls) using generalized linear mixed models with the adjustment of age, sex, population structure, genomic relationship matrix, and genotyping batches. Next, using methylation and bulk RNA-sequencing data from the dorsolateral pre-frontal cortex in 150 Hispanics brains, we tested the cis- and trans-effects of AD associated CGS on brain DNA methylation to mRNA expression. For the genes with significant cis- and trans-effects, we checked their enriched pathways.

**Results:** We identified six genetic loci in Hispanics with CGS dosage associated with AD at genome-wide significance levels: *ADAM20* (Score=55.2, *P=*4.06×10^-8^), between *VRTN* (Score=-19.6, *P=*1.47×10^-8^) and *SYNDIG1L* (Score=-37.7, *P=*2.25×10^-9^), *SPG7* (16q24.3) (Score=40.5, *P=*2.23×10^-8^), *PVRL2* (Score=125.86, *P=*1.64×10^-9^), *TOMM40* (Score=-18.58, *P=*4.61×10^-8^), and *APOE* (Score=75.12, *P=*7.26×10^-26^). CGSes in *PVRL2* and *APOE* were also genome-wide significant in NHW. Except for *ADAM20*, CGSes in all the other five loci were associated with Hispanic brain methylation levels (mQTLs) and CGSes in *SPG7, PVRL2,* and *APOE* were also mQTLs in NHW. Except for *SYNDIG1L* (*P*=0.08), brain methylation levels in all the other five loci affected downstream RNA expression in the Hispanics (*P*<0.05), and methylation at *VRTN* and *TOMM40* were also associated with RNA expression in NHW. Gene expression in these six loci were also regulated by CpG sites in genes that were enriched in the neuron projection and synapse (FDR<0.05).

**Conclusions:** We identified six CpG associated genetic loci associated with AD in Hispanics, harboring both genetic and epigenetic risks. However, their downstream effects on mRNA expression maybe ethnic specific and different from NHW.

## Introduction

Alzheimer disease (AD) is a chronic and progressive neurodegenerative disorder accompanied by cognitive decline that gradually worsen over years. The etiology of AD is complex involving different molecular mechanisms, which may be the result of not only heritable genetic risks but also by factors that act on the epigenome. The advancement in identifying genetic contributions to AD has also piqued interest in epigenetic contributions. The most recent genome-wide association study (GWAS) of AD reported over 70 genetic loci for AD risk[1]. Candidate gene and genome-wide DNA methylation studies have implicated approximately 21 genetic loci with differential methylation levels associated with AD[2].

Loci identified in both genetic and epigenetic studies[3] suggest a common molecular hub that captures causal risk factors for AD. *APOE* ε4 is most consistently confirmed genetic risk factor for AD and the methylation levels of the CpG island within *APOE* were found to be lower in AD brains compared to brains from healthy participants[4]. The study of CpG-related single nucleotide polymorphism (CGS) may identify alleles disrupting the CpG dinucleotides leading to the removal of the DNA methylation targeted site. We previously found CGSes in the MS4A region have a dose-dependent effect on AD in persons who identify as non-Hispanic White (NHW) [3]. We identified a statistical association between MS4A CGSes and DNA methylation levels using blood samples and clinical AD but we and others were limited in the downstream functional validation for the top loci.

In this study, we conducted a systematic analysis based on CGS in persons who identified as Hispanics who enrolled Washington Heights-Inwood Columbia Aging Project (WHICAP) or the Estudio Familiar de Influencia Genética en Alzheimer (EFIGA)[5]. We have studied AD in numerous cohorts of persons who have identified as Hispanics and understand their complex ancestry. We began by using a genome-wide sliding window approach to prioritize genetic loci comprised of CGSes associated with the risk of clinical diagnosis of AD in 7,155 Hispanics decedents. The prioritized loci from the genome-wide analyses were followed by detailed functional studies. We analyzed both the cis- and trans-effects of the molecular mechanisms from genetics to DNA methylation and mRNA gene expression in postmortem brain tissue from Hispanics decedents.

## Methods

### Study description

#### Cohorts included for genetic studies

We included 7,155 Hispanics decedents from the Washington Heights-Inwood Columbia Aging Project (WHICAP)[6] and the Estudio Familiar de Influencia Genética en Alzheimer (EFIGA)[7]. The WHICAP study is an ongoing prospective, community-based, multiethnic longitudinal study of Medicare beneficiaries 65 years and older residing in northern Manhattan (Washington Heights, Hamilton Heights, and Inwood). All the participants underwent a comprehensive examination including the assessment of general health and function, standardized physical and neurological examination, and a neuropsychological battery of tests. Follow-up visits were performed every 1.5-2 years, repeating similar examinations. Initiated in 1998, the EFIGA recruited individuals of Caribbean Hispanic ancestry including familial and sporadic AD. The individuals were recruited in New York City using local newspapers, the local Caribbean Hispanic radio station, and postings throughout the Washington Heights-Inwood neighborhood. AD was defined as any individual meeting NINCDS-ADRDA criteria for probable or possible AD[8]. The severity of dementia was rated according to the Clinical Dementia Rating[9].

We also included 1,283 NHW from the Religious Order Study and the Memory & Aging Project (ROSMAP) study to analyze whether the top loci identified in the Hispanics can be generalized to NHW. ROSMAP recruit older individuals without known dementia and their detailed information of both ante-mortem and post-mortem phenotyping were collected[10]. For this study, we have included in total 1,283 NHW with whole genome sequencing data and clinical diagnosis of AD.

#### Cohorts with brain DNA methylation and RNA sequencing (RNA-seq)

New York Brain Bank (NYBB): Brain tissue came from The NIA Alzheimer’s disease Family Based Study (NIA-AD FBS), WHICAP, EFIGA, and NCRAD. The NIA-AD FBS included 9,682 family members, and 1,096 unrelated, nondemented elderly from different race/ethnicity groups from 1,756 families with suspected AD. NCRAD included unaffected individuals from families with a history of AD. A description of the families has been previously detailed in a report [5].

University of California, Davis Alzheimer’s Disease Center (UCD ADC): With the aim to conduct a research on diversity and risk of AD dementia, UCD applied an active community outreach approach to recruit the individuals from the communities of Alameda, Contra Costa, Sacramento, San Joaquin, Solano, and Yolo County. The overall percentage of Hispanic individual 60 years of age and older residing in these counties ranged from 7.1% to 14.3% [11].

Florida Autopsied Multi-Ethnic (FLAME) cohort: The FLAME cohort is derived from the State of Florida brain bank housed at the Mayo Clinic Florida [12]. The FLAME cohort consists of a total of 2,809 autopsied individuals with a wide range of neurodegenerative diseases, who were self-identified as Hispanic/Latino, black/African, and non-Hispanic white/European. The fixed hemi-brain (typically left hemisphere) was weighed and the frontal cortex was cut and then placed in 10% formalin solution.

The University of Pennsylvania Integrated Neurodegenerative Disease Biobank: Patients with neurodegenerative disease are recruited into the autopsy program by the different clinical cores. The subjects selected for the autopsy were followed in the clinical centers with detailed clinical information and most of them were also collected with biofluid, neuroimaging, and genetic data/samples. The left hemisphere and brain stem were immersed in 10% neutral buffered formalin for 2 weeks whereas the right hemisphere was sliced coronally and frozen.

University of California, San Diego Alzheimer’s Disease Research Center (UCSD ADC): Postmortem tissue from the center’s longitudinally followed cohort was used for this study. Blocks of tissue were provided from autopsy verified cases after fixation in 10% formalin for 4 weeks. Cases were selected by using detailed clinical, biomarker and demographics information collected at visits.

The Religious Order Study and the Memory & Aging Project (ROSMAP): We have included 516 NHW who have measurements of both postmortem brain DNA methylation and RNA sequencing (RNA-seq) from their postmortem brain tissues. The details of both datasets were described previously [10]. In brief, the grey matter from the dorsolateral prefrontal cortex (DLPFC) were dissected while still frozen. RNA was extracted for transcriptome library construction following the dUTP protocol and Illumina sequencing. The extracted DNA were processed on the Illumina Infinium HumanMethylation450 BeadChip.

An informed consent was signed by the participant and/or legal guardian of the individuals included into this study. IRB approval was approved by each institution.

### Genotype data and annotations of CpG-related SNPs (CGS)

We annotated the CpG-related single nucleotide polymorphisms (CGS) by checking the three base pairs flanking each single nucleotide polymorphism (SNP) and derived the dosage of the CpG site-creating allele of multiple CGSes in each 1 Kb window across the genome. We imputed the genotype according to Haplotype Reference Consortium (HRC) reference panels in 7,155 Caribbean Hispanics. We used the whole genome sequencing data of 1,283 NHW.

### Brain DNA methylation data

The genome-wide DNA methylation profile was measured by the Infinium MethylationEPIC Kit (Illumina). For each sample we checked the control probes, sex mismatches, contamination, and genotype outliers calling to identify and remove those samples failed quality control (QC). We kept those CpG sites with detection *P* value < 0.01 across all the qualified samples and masked those sample-specific CpG site with new detection *P* > 0.01 [13]. We further removed those CpG sites reported to have cross-hybridization problems [14, 15] and those polymorphic CpG sites [15, 16]. We further corrected the dye bias for all the qualified CpG probes. Finally, there were 179 samples from person of Hispanic descent with 675,583 autosomal probes passed the QC which were included into the current study.

Brain RNA-seq data: Total RNA was extracted using Qiagen’s RNeasy Mini Kit and then was sent to the New York Genome Center for transcriptome library construction, which was sequenced on a NovaSeq 6000 flow cell using 2×100bp cycles, targeting 60 million reads per sample. All the samples included into the analysis passed QC metrics using *FastQC*. Gene counts were calculated using the function *featureCounts*. We applied *ComBat-seq* to correct batch effects. As a result, a total of 58,942 genes passed QC metrics and exhibiting non-zero expression across all participants.

### Statistical analysis

We scaled the dosage of each window to fit into the value from zero to 2 and then tested the scaled dosage of each window for association with AD using generalized linear mixed models (GLMMs) implemented in the generalized linear mixed model association tests (GMMAT) [17] with the adjustments for age, sex, population substructure, genomic relationship matrix (GRM), and genotyping batches. Genome-wide significance threshold was *p* <5.0×10^-8^. For the mQTL analysis, there were 112 samples from persons of Hispanic decent and 571 from persons of NHW decent with both genotype and brain DNA methylation data. We conducted the generalized linear model adjusting for the age at death, sex, and technical covariates of genotyping and methylation batches, and methylation chip and position. We analyzed both the cis- and trans-effect of the DNA methylation on gene expression. For the cis-effect of DNA methylation on gene expression, we conducted a highly adaptive sum of powered score-weighted test (aSPUw) to collapse all the available CpG sites within 100 Kb distance to the gene (from 50 Kb upstream of the transcription start site and 50 Kb downstream of the end site of the gene according to GENCODE v44 (GRCh37) annotation) with the adjustment of age, sex, and technical covariates of methylation chip ID and chip position. For the trans-effect of DNA methylation on gene expression, we used the linear mixed model to control for the random effect of methylation array, the fixed covariates of chip position on the methylation array, the batch effects, age at death, and sex.

### Protein-protein interactive network and pathway analysis

There were 69 CpG sites annotated to 65 genes associated with the mRNA expression level in human brains. We searched their network enrichment and pathway analysis through STRING (https://string-db.org).

## Results

### Characteristics of the individuals providing brain samples

There were brain samples from 179 Hispanics and 571 NHW individuals included in the study, and their demographics are shown in **Table 1**. The mean age at death for Hispanics is 80 years while it is 88 years for NHW individuals. 57.54% of Hispanics and 62.9% of NHW were women.

**Table 1.** Characteristics of donors of brain tissues.

|  | Hispanics (N=179) |  |  | Non-Hispanic Whites (N=571) |
| --- | --- | --- | --- | --- |
|  | All (N=179) | With GWAS (N=112) | With RNA-seq (N=150) |  |
| Age at death (y)* | 80.29 (11.33) | 78.52 (11.24) | 81.08 (10.34) | 88.32 (6.52) |
| Female (N)<sup>#</sup> | 103 (57.54%) | 64 (57.14%) | 83 (55.33%) | 359 (62.9%) |

\*The mean and standard deviation of the age at death are shown.
<sup>#</sup>The number and percentage of female are shown.

### CpG identification

Within the 7,155 Hispanic participants, 1,857,611 windows of 1 Kb genome-wide, with at least two CpG sites, were tested for association with AD. Using the Bonferroni corrected genome-wide significance p-value of <5.0×10^-8^, we identified six genome-wide significant regions: *ADAM20* (Score=55.2, *P=*4.06×10^-8^), between *VRTN* (Score=-19.6, *P=*1.47×10^-8^) and *SYNDIG1L* (Score=-37.7, *P=*2.25×10^-9^), *SPG7* (16q24.3) (Score=40.5, *P=*2.23×10^-8^), *PVRL2* (Score=125.86, *P=*1.64×10^-9^), *TOMM40* (Score=-18.58, *P=*4.61×10^-8^), and *APOE* (Score=75.12, *P=*7.26×10^-26^). (**Figure 1 & Table 2**). The CGS windows in *PVRL2* (Score=11.25, *P=*2.4×10^-6^) and *APOE* (Score=14.46, *P=*1.53×10^-11^) were also significant in the 1,283 NHW participants from ROSMAP.

**Figure 1.**
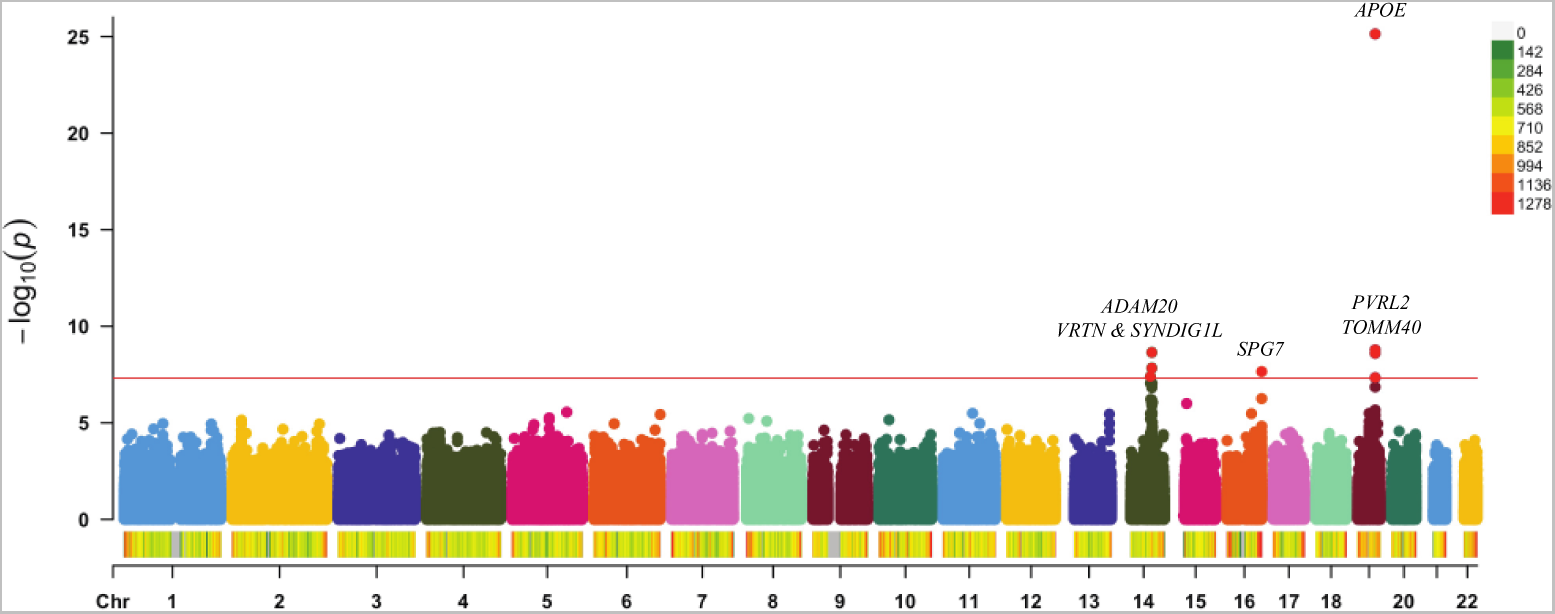
Manhattan plot of sliding window search across the genome for the risk loci of clinical diagnosis of Alzheimer disease. Each dot represents one 1-Kb window, and X and Y axis shows its genomic coordinate and -log10 transformed *P* value. The horizontal black line shows the Bonferroni-corrected genome-wide significance threshold (*P*≤ 5×10^-8^). The density of the windows across the genome were shown as color coded bars with the color coding is shown on the right.

**Table 2.** Top CGS windows associated with Alzheimer’s disease.

| Chr | Window Position | Gene | Hispanics in WHICAP (N=7155) |  |  | Whites in ROSMAP (N=1283) |  |  |
| --- | --- | --- | --- | --- | --- | --- | --- | --- |
|  |  |  | SCORE | VAR | P | SCORE | VAR | P |
| 14 | chr14:70994207-70995206 | *ADAM20* | 55.19 | 101.12 | 4.06E-08 | 1.06 | 1.66 | 0.41 |
| 14 | chr14:74867207-74868206 | *VRTN* and *SYNDIG1L* | -37.67 | 39.70 | 2.25E-09 | -0.20 | 0.11 | 0.56 |
| 16 | chr16:89588052-89589051 | *SPG7* | 40.51 | 52.45 | 2.23E-08 | -3.06 | 3.51 | 0.10 |
| 19 | chr19:45387308-45388307 | *PVRL2* | 125.86 | 435.69 | 1.64E-09 | 11.25 | 5.69 | 2.40E-06 |
| 19 | chr19:45402808-45403807 | *TOMM40* | -18.58 | 11.56 | 4.61E-08 | -0.13 | 0.009 | 0.17 |
| 19 | chr19:45411308-45412307 | *APOE* | 75.12 | 51.03 | 7.26E-26 | 14.46 | 4.60 | 1.53E-11 |

The dosage of CpG dinucleotides created by multiple CpG-related single nucleotide polymorphisms (CGSs) of each window were scaled into the value from 0 to 2, which was analyzed for its association with Alzheimer's disease using generalized linear mixed models (GLMMs) implemented in the generalized linear mixed model association tests (GMMAT) with the adjustment of age, sex, and genotyping batches with the random effects of both kinship and genomic relationship matrix (GRM).

### Cis-effects of CGS on DNA methylation

We tested the cis-effects of CGS on molecular phenotypes within 50 Kb flanking the gene. The cis-effects on DNA methylation of the CpG dosage of several CGSes in the windows of AD are presented in **Table 3**. Except for *ADAM20*, all the other five loci have significant associations between the CGSes dosage and the DNA methylation level of the CpG sites within the cis-regions: the intergenic region between *VRTN* and *SYNDIG1L* (cg16837088, b=-0.04, *P*=2.94×10^-3^), *SPG7* (cg26536240, b=0.02, *P*=5.78×10^-7^), *PVRL2* (cg04406254, b=0.02, *P*=2.49×10^-3^), *TOMM40* (cg20051876, b=-0.04, *P*=0.02), and *APOE* (cg20090143, b=-0.01, *P*=0.02). In NHW, different CpG sites showed statistical significance at *SPG7* (cg02244288, b=-0.01, *P*=2.76×10^-63^), *PVRL2* (cg02613937, b=-0.01, *P*=2.27×10^-6^), and *APOE* (cg02613937, b=-0.01, *P*=2.36×10^-6^). The cis-mQTLs at *ADAM20* (cg04910453, b=-0.02, *P*=0.05) reached nominal significance (*P*≤0.05) in NHW but not in Hispanics.

**Table 3.**
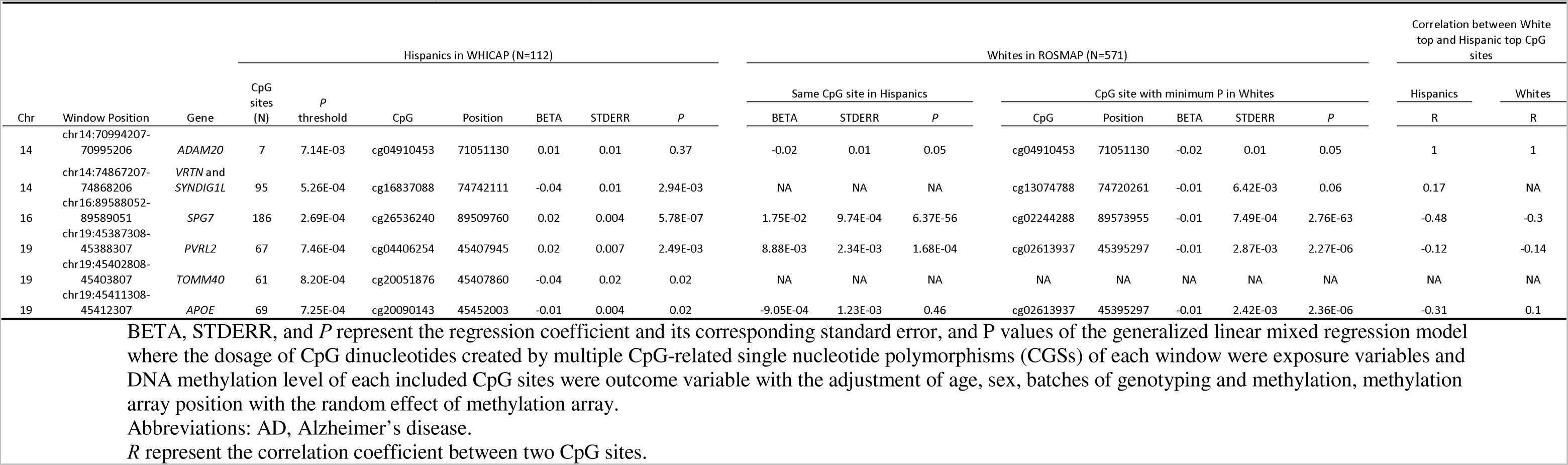
Cis-effects of CGS dosage on DNA methylation.

### DNA methylation levels altering downstream cis-mRNA expression

Next, we tested the methylation sites cis-regulated by AD-associated CGSes (identified above), to determine whether these sites altered downstream mRNA expression in the brain. Because our findings revealed different methylation sites for the same gene in Hispanics and NHW for several AD associated loci, we conducted an aggregate analysis by collapsing all the CpG sites within the cis-region of the targeted gene (**Table 4**). We found that except for *SYNDIG1L*, methylation levels in all the other genes significantly altered the brain RNA expression in Hispanics (*P*≤0.05). The significance was replicated for *VRTN* and *TOMM40* in NHW (*P*≤0.05).

**Table 4.** Cis-effects of DNA methylation on gene expression.

| Chr | Gene | Hispanics in WHICAP (N=112) |  |  |  |  | Whites in ROSMAP (N=516) |  |  |  |  |
| --- | --- | --- | --- | --- | --- | --- | --- | --- | --- | --- | --- |
|  |  | SPUw1 |  | aSPUw |  | P values range | SPUw1 |  | aSPUw |  | P values range |
|  |  | T | P | T | P |  | T | P | T | P |  |
| 14 | *ADAM20* | -7.25 | 0.09 | 0.01 | 0.03 | [0.012, 0.09] | 4.04E-16 | 0.16 | 0.10 | 0.25 | [0.1, 0.55] |
| 14 | *VRTN* | -63.76 | 0.05 | 0.003 | 8.00E-03 | [0.003, 0.046] | 9.99E-16 | 0 | 0 | 9.99E-04 | [0, 0.000999] |
| 14 | *SYNDIG1L* | -26.52 | 0.40 | 0.03 | 0.08 | [0.03, 0.40] | 1.09E-14 | 0.25 | 0.03 | 0.08 | [0.03, 0.25] |
| 16 | *SPG7* | -129.93 | 0.002 | 0 | 9.99E-04 | [0, 0.005] | 5.01E-12 | 0.47 | 0.27 | 0.61 | [0.27, 0.62] |
| 19 | *PVRL2* | -75.43 | 0.002 | 0 | 9.99E-04 | [0, 0.002] | -4.57E-13 | 0.19 | 0.12 | 0.25 | [0.12, 0.25] |
| 19 | *TOMM40* | 17.37 | 0.38 | 0.003 | 0.01 | [0.003, 0.38] | 8.57E-13 | 0.02 | 0.02 | 0.03 | [0.02, 0.03] |
| 19 | *APOE* | -83.13 | 0.001 | 0 | 9.99E-04 | [0, 0.001] | -2.07E-11 | 0.96 | 0.04 | 0.12 | [0.04, 0.97] |

### Trans-effects of DNA methylation levels on gene expression

We tested whether the expression of the genes that harbor AD associated CGS were influenced by genome wide CpG sites in trans. We identified 69 CpG sites across the genome that regulated gene expression of *ADAM20*, *SYNDIG1L*, *SPG7*, *PVRL2*, *TOMM40*, and *APOE* in Hispanics at genome-wide significant levels after Bonferroni correction of the number of CpG sites included into the analysis (**Supplementary Table 2**). At *PVRL2* and *TOMM40*, the same CpG sites regulating the gene expression in Hispanics also regulated the gene expression in NHW (P<0.05).

These 69 CpG sites which have the trans-effects on the gene expressions were annotated to 65 genes. We combined these 65 genes with the target genes of *ADAM20*, *SYNDIG1L*, *SPG7*, *PVRL2*, *TOMM40*, and *APOE*, to be upload to STRINGdb to query the significant biological pathways. The significant pathways (FDR<0.05) were presented in **Table 5**, which involved neuron projection and glutamatergic synapse (FDR=0.0189).

**Table 5.** Pathway analysis of the trans-effects of DNA methylation on gene expression.

| Category | Term ID | Term Description | Observed Gene Count | Background Gene Count | Strength | False Discovery Rate |
| --- | --- | --- | --- | --- | --- | --- |
| GO Component | GO:0030054 | Cell junction | 19 | 2115 | 0.43 | 0.0189 |
| GO Component | GO:0030424 | Axon | 11 | 651 | 0.71 | 0.0189 |
| GO Component | GO:0043005 | Neuron projection | 16 | 1391 | 0.54 | 0.0189 |
| GO Component | GO:0098978 | Glutamatergic synapse | 8 | 334 | 0.86 | 0.0189 |
| Monarch | EFO:0004612 | High density lipoprotein cholesterol measurement | 14 | 740 | 0.76 | 0.0015 |
| Monarch | EFO:0004732 | Lipoprotein measurement | 17 | 1426 | 0.56 | 0.017 |
| Monarch | EFO:0004614 | Apolipoprotein A 1 measurement | 9 | 396 | 0.84 | 0.0275 |
| Monarch | EFO:0005105 | Lipid or lipoprotein measurement | 22 | 2526 | 0.42 | 0.0345 |
| Monarch | EFO:0004529 | Lipid measurement | 21 | 2400 | 0.42 | 0.0365 |
| Monarch | EFO:0004582 | Liver enzyme measurement | 14 | 1124 | 0.58 | 0.0365 |
| Monarch | EFO:0004747 | Protein measurement | 36 | 5856 | 0.27 | 0.0365 |
| TISSUES | BTO:0001484 | Nervous system | 42 | 6016 | 0.33 | 4.03e-05 |
| TISSUES | BTO:0000227 | Central nervous system | 39 | 5825 | 0.31 | 0.00044 |
| TISSUES | BTO:0000142 | Brain | 38 | 5733 | 0.3 | 0.00067 |
| TISSUES | BTO:0000282 | Head | 39 | 6642 | 0.25 | 0.0081 |
| COMPARTMENTS | GOCC:0030054 | Cell junction | 15 | 1053 | 0.64 | 0.0033 |
| COMPARTMENTS | GOCC:0045202 | Synapse | 9 | 493 | 0.74 | 0.0426 |
| UniProt Keywords | KW-0025 | Alternative splicing | 52 | 10313 | 0.18 | 0.0024 |

## Discussion

We have conducted a first multi-omics investigation of CpG related SNPs (CGS) in brain tissue from a group of individuals of Hispanic descent, which confer both genetic and epigenetic effects among individuals. Our study is one of the largest genome-wide association studies with the focus on the special type of genetic variants of CGS in Hispanics. We then assessed the effect cascade from the AD-associated CGSes to brain methylation levels to brain expression levels. This study was unique in terms of its use of Hispanic human brain tissues for AD. Also, the current study provided robust results which survived the most stringent Bonferroni corrections on the multiple testing.

We identified six genome-wide significant windows in or near *ADAM20*, *VRTN*, *SYNDIG1L*, *SPG7*, *PVRL2*, *TOMM40*, and *APOE*, where the dosage of the CpG dinucleotides (created by the including CGSes) were associated with the risk of clinical diagnosis of AD. In *SPG7,* the AD associated CGS window is associated with increased cis-DNA methylation levels in the frontal cortex, which in turn reduced downstream mRNA expression. We validated the *SPG7* genetic and epigenetic alterations in NHW, but there is no effect on mRNA expression. Similarly, for *SYNDIG1L* and *APOE,* we identified AD-associated CGSes which in turn regulated methylation levels in both Hispanics and NHW brains. However, but the epigenetic modifications had muted effect on downstream gene expression. At *PVRL2*, both the cis effects and trans-effects were statistically significant.

*SPG7* gene encodes paraplegin, a component of the m-AAA protease, an ATP-dependent proteolytic complex of the mitochondrial inner membrane that degrades misfolded proteins and regulates ribosome assembly. Our finding of its significant effects of its association with AD was consistent with the previous report that the DNA methylation level at SPG7 was associated with Braak neurofibraillary stages[18]. The *PVRL2* (a.k.a. *NECTIN2*) gene encodes a gene within the nectin subfamily of immunoglobulin-like adhesion molecules that participate in Ca^2+^-independent cell-cell adhesion. It is upstream of *TOMM40* and *APOE* and located within the highly linked genetic cluster of *TOMM40*-*APOE*-*APOC2*. *PVRL2* had both cis- and trans-effects between DNA methylation and mRNA gene expression. The CpG island within *APOE* was reported to have lower DNA methylation level in AD patients compared to controls in human postmortem brains[19, 20], which is more profound in glial cells[19]. Lee[21] reported a negative correlation between *APOE* total RNA levels and DNA methylation at the CpG island within *APOE* in human postmortem frontal lobes, and this negative correlation is more obvious in controls compared to AD patients. *SYNDIG1L* (also known as *TMEM90A* or *CAPUCIN*) encodes synapse differentiation-induced gene 1 like. In rodents, memory and motor deficits caused by 1,2-Diacetylbenzene via alteration of the mRNA expression of *Syndig1l* [22] can be improved by prolactin.

Although this project is currently the largest one with Hispanic brain DNA methylation data, it does have limitations of potential bias by grey vs. white matter composition driven by different protocol used by different sites. Also, the fact that multiple sites contribute to the brain samples may also bring variations into the findings. Since the brain samples from different sites were measured by different methylation batches, we only adjust for the experiment batches not the sites in the regression model to remove the colinear bias.

This is the first report of robust six genetic loci covering seven genes that act as the hub for both the genetic and epigenetic effects on clinical diagnosis of AD in Hispanic: *ADAM20*, between *VRTN* and *SYNDIG1L*, *SPG7*, *PVRL2*, *TOMM40*, and *APOE*. *PVRL2* and *APOE* were also genetically significant in NHW. Except *ADAM20*, all the other loci have significant mQTL effects in Hispanics, and *SPG7*, *PVRL2*, *APOE* also have significant mQTL in NHW. The DNA methylation levels of all seven genes except for *SYNDIG1L* have significant associations with its mRNA gene expression levels in Hispanic brains, while only *VRTN* and *TOMM40* also showed significant associations on mRNA expression levels in NHW brains. Except for *VRTN*, the mRNA gene expression levels of all the other six genes have significant trans-effects from DNA methylation levels of the CpG sites in Hispanics, while only *PVRL2* and *TOMM40* also showed trans-effects in NHW.

We have identified in total six genetic loci*PVRL2* had both significant cis- and trans-effects from the genetics to epigenetics and then to the mRNA gene expression and the genes for the trans-effects are enriched in the pathways of neuron projection and glutamatergic synapse. Except for *SYNDIG1L SPG7* and *APOE* had significant cis-effects while *SYNDIG1L* has significant trans-effects.

## Data Availability

All data produced in the present study are available upon reasonable request to the authors

## Acknowledgements

We thank all the investigators for this study. We thank Dr. David Bennett for releasing ROSMAP datasets publicly. Data collection and sharing for this project was supported by (WHICAP, R01 AG072474, RF1 AG066107) funded by the National Institute on Aging (NIA) and by the National Center for Advancing Translational Sciences, National Institutes of Health, through Grant Number UL1TR001873. Data collection for this project was supported by the Genetic Studies of Alzheimer disease in Caribbean Hispanics (EFIGA) funded by the National Institute on Aging (NIA) and by the National Institutes of Health (NIH) (R01 AG067501). The project was partially funded by the National Institute on Aging (NIA) of the National Institutes of Health (NIH) under Award Numbers R01AG062517, and P30AG072972, P30AG062429, NIA (P30 AG062677, P01 AG003949), Florida Department of Health, Ed and Ethel Moore Alzheimer Disease Research Program (20A22, 8AZ06), NIH P30AG072979, P01AG066597 and U19AG062418.

