## Supplemental Tables for "Multi-omics Characterization of Epigenetic and Genetic Risk of Alzheimer Disease in Autopsied Brains from two Ethnic Groups"

**Supplementary Tables**

Supplementary Table 1. Characteristics of the total of 179 Hispanic brain donors stratified by different studies.

|  | With GWAS (N=112) | | |  | With RNAseq (N=150) | | |
| --- | --- | --- | --- | --- | --- | --- | --- |
| Study | N | Age at death* (y) | Female^#^ (N) |  | N | Age at death* (y) | Female^#^ (N) |
| NYBB | 20 | 81.80 (8.57) | 16 (80%) |  | 42 | 82.46 (10.74) | 28 (66.67%) |
| MAYO | 79 | 77.84 (11.11) | 40 (50.63%) |  | 59 | 79.22 (8.79) | 29 (49.15%) |
| UCDavis | 6 | 82.00 (13.33) | 4 (66.67%) |  | 3 | 87.33 (4.73) | 1 (33.33%) |
| Upenn | 7 | 73.86 (16.56) | 4 (57.14%) |  | 7 | 73.86 (16.56) | 4 (57.14%) |
| UCSD | 0 | NA | NA |  | 39 | 83.21 (10.37) | 21 (53.85%) |

*The mean and standard deviation of the age at death are shown.

^#^The number and percentage of female are shown.

Supplementary Table 2. Top CpG sites for their trans-effects on the mRNA expression of *SYNDIG1L*, *APOE*, and *PVRL2*.

|  |  |  |  |  | Hispanics in WHICAP (N=150) | | |  | Whites in ROSMAP (N=516) | | |
| --- | --- | --- | --- | --- | --- | --- | --- | --- | --- | --- | --- |
| Targeted Gene | Chr | BP | CpG site | Gene of CpG site | BETA* | STDERR* | *P** |  | BETA* | STDERR* | *P** |
| *ADAM20* | 17 | 38504102 | cg19709355 | *RARA* | 883 | 151 | 6.45e-08 |  | 2.22 | 4.16 | 0.59 |
| *ADAM20* | 17 | 79047823 | cg25043129 | *BAIAP2* | -713 | 122 | 7.1e-08 |  | 3.65 | 3.14 | 0.25 |
| *SYNDIGL1* | 1 | 1243747 | cg25201327 | *ACAP3* | 1640 | 277 | 5.13e-08 |  | NA | NA | NA |
| *SYNDIGL1* | 13 | 47371311 | cg04277327 | *ESD* | 838 | 140 | 3.43e-08 |  | -103.00 | 62.60 | 0.10 |
| *SPG7* | 3 | 40566228 | cg17545383 | *ZNF621* | 43900 | 7530 | 7.53e-08 |  | 334.00 | 2900.00 | 0.91 |
| *PVRL2* | 1 | 3784725 | cg03591123 | *DFFB* | -4120 | 658 | 1.08e-08 |  | NA | NA | NA |
| *PVRL2* | 1 | 6266416 | cg11764177 | *RNF207* | -3470 | 601 | 9.48e-08 |  | 225.00 | 287.00 | 0.43 |
| *PVRL2* | 1 | 21901166 | cg07142010 | *ALPL* | -2570 | 433 | 4.71e-08 |  | 128.00 | 197.00 | 0.52 |
| *PVRL2* | 1 | 36773099 | cg15131207 | *SH3D21* | 4180 | 713 | 6.15e-08 |  | NA | NA | NA |
| *PVRL2* | 1 | 43534145 | cg18153322 | *RNU6-870P* | -4710 | 764 | 1.66e-08 |  | 183.00 | 419.00 | 0.66 |
| *PVRL2* | 1 | 45273495 | cg24254387 | *lncRNA_ENSG00000290041* | -2440 | 416 | 6.61e-08 |  | 171.00 | 221.00 | 0.44 |
| *PVRL2* | 1 | 233585348 | cg12166740 | *RNU4-77P* | -5050 | 814 | 1.41e-08 |  | NA | NA | NA |
| *PVRL2* | 2 | 75059030 | cg08729839 | *HK2* | -4140 | 649 | 6.35e-09 |  | -17.40 | 330.00 | 0.96 |
| *PVRL2* | 2 | 182678354 | cg17558303 | *RNU6ATAC19P_ITPRID2-D* | -4570 | 697 | 2.85e-09 |  | NA | NA | NA |
| *PVRL2* | 2 | 241385598 | cg26128123 | *GPC1* | -3450 | 518 | 1.65e-09 |  | 461.00 | 378.00 | 0.22 |
| *PVRL2* | 4 | 52905025 | cg12489960 | *SGCB* | 3040 | 503 | 3.01e-08 |  | -113.00 | 336.00 | 0.74 |
| *PVRL2* | 4 | 57604055 | cg02576414 | *RPL17P20* | -3900 | 599 | 3.41e-09 |  | NA | NA | NA |
| *PVRL2* | 4 | 154047992 | cg00554773 | *TRIM2* | -4600 | 700 | 2.71e-09 |  | NA | NA | NA |
| *PVRL2* | 5 | 150029342 | cg18416096 | *SYNPO* | 4090 | 571 | 1.54e-10 |  | -213.00 | 376.00 | 0.57 |
| *PVRL2* | 5 | 150385881 | cg15550277 | *GPX3* | -2900 | 492 | 5.68e-08 |  | NA | NA | NA |
| *PVRL2* | 5 | 175961105 | cg26405376 | *RNF44* | -2910 | 483 | 3.11e-08 |  | 90.10 | 306.00 | 0.77 |
| *PVRL2* | 5 | 177679652 | cg21948676 | *COL23A1* | -4180 | 675 | 1.44e-08 |  | NA | NA | NA |
| *PVRL2* | 6 | 31747041 | cg26137417 | *VARS1* | -3230 | 446 | 1.07e-10 |  | 269.00 | 272.00 | 0.32 |
| *PVRL2* | 6 | 32016236 | cg19964491 | *TNXB* | -2250 | 342 | 2.53e-09 |  | -165.00 | 212.00 | 0.44 |
| *PVRL2* | 6 | 32016239 | cg11493661 | *TNXB* | -2430 | 347 | 3.68e-10 |  | -335.00 | 202.00 | 0.10 |
| *PVRL2* | 7 | 1100172 | cg00738919 | *C7ORF50* | -3160 | 536 | 5.55e-08 |  | -278.00 | 280.00 | 0.32 |
| *PVRL2* | 7 | 4809628 | cg26738657 | *FOXK1* | -3700 | 531 | 4.02e-10 |  | NA | NA | NA |
| *PVRL2* | 7 | 4869981 | cg00469380 | *RADIL* | -2280 | 383 | 4.41e-08 |  | -231.00 | 163.00 | 0.16 |
| *PVRL2* | 8 | 29914060 | cg26836824 | *SARAF* | -3740 | 644 | 8.27e-08 |  | NA | NA | NA |
| *PVRL2* | 8 | 87688230 | cg11689702 | *CNGB3* | -4680 | 796 | 5.85e-08 |  | NA | NA | NA |
| *PVRL2* | 9 | 132195129 | cg17977699 | *lncRNA_ENSG00000230676* | -5240 | 816 | 5.09e-09 |  | NA | NA | NA |
| *PVRL2* | 9 | 135997155 | cg16872071 | *RALGDS* | 3450 | 571 | 3.02e-08 |  | -47.10 | 313.00 | 0.88 |
| *PVRL2* | 10 | 87912236 | cg21605505 | *GRID1* | -3910 | 671 | 7.41e-08 |  | NA | NA | NA |
| *PVRL2* | 11 | 35308945 | cg24175498 | *SLC1A2* | -2970 | 514 | 9.44e-08 |  | NA | NA | NA |
| *PVRL2* | 11 | 47290613 | cg26365553 | *NR1H3* | 3160 | 526 | 3.27e-08 |  | -434.00 | 287.00 | 0.13 |
| *PVRL2* | 11 | 65343330 | cg15217978 | *lncRNA_ENSG00000289339* | -4280 | 665 | 4.76e-09 |  | 277.00 | 327.00 | 0.40 |
| *PVRL2* | 11 | 67070738 | cg16802508 | *SSH3* | -2930 | 507 | 9.45e-08 |  | -213.00 | 270.00 | 0.43 |
| *PVRL2* | 11 | 117314759 | cg09519504 | *DSCAML1* | -3890 | 661 | 6.02e-08 |  | NA | NA | NA |
| *PVRL2* | 12 | 58211193 | cg01424889 | *AVIL* | -3670 | 558 | 2.51e-09 |  | 198.00 | 215.00 | 0.36 |
| *PVRL2* | 12 | 133083102 | cg07578029 | *FBRSL1* | -3140 | 483 | 3.52e-09 |  | NA | NA | NA |
| *PVRL2* | 14 | 104178543 | cg11718757 | *XRCC3* | -3720 | 471 | 4.69e-12 |  | NA | NA | NA |
| *PVRL2* | 14 | 105942243 | cg08182193 | *CRIP2* | -2380 | 408 | 7.51e-08 |  | -160.00 | 290.00 | 0.58 |
| *PVRL2* | 16 | 2083393 | cg08601673 | *SLC9A3R2* | -1950 | 308 | 7.16e-09 |  | 543.00 | 265.00 | 0.04 |
| *PVRL2* | 16 | 75286915 | cg05389731 | *BCAR1* | -2780 | 468 | 4.41e-08 |  | -59.90 | 267.00 | 0.82 |
| *PVRL2* | 17 | 42989137 | cg20911989 | *GFAP* | -2870 | 497 | 9.36e-08 |  | -40.60 | 236.00 | 0.86 |
| *PVRL2* | 17 | 73471255 | cg08055304 | *TMEM94* | -3580 | 593 | 2.86e-08 |  | NA | NA | NA |
| *PVRL2* | 17 | 73726430 | cg13347255 | *ITGB4* | -4180 | 611 | 7.41e-10 |  | -10.40 | 485.00 | 0.98 |
| *PVRL2* | 17 | 80241424 | cg01194390 | *lncRNA_ENSG00000287737* | 3580 | 563 | 6.68e-09 |  | NA | NA | NA |
| *PVRL2* | 19 | 1155030 | cg19649900 | *SBNO2* | -2420 | 389 | 1.24e-08 |  | -160.00 | 274.00 | 0.56 |
| *PVRL2* | 19 | 19416751 | cg01313994 | *SUGP1* | 3260 | 558 | 6.93e-08 |  | 171.00 | 272.00 | 0.53 |
| *PVRL2* | 20 | 31127002 | cg08006263 | *NOL4L* | 3250 | 503 | 4.41e-09 |  | NA | NA | NA |
| *TOMM40* | 2 | 242170211 | cg09235936 | *HDLBP* | -1380 | 219 | 9.26e-09 |  | 871.00 | 368.00 | 0.02 |
| *TOMM40* | 2 | 242170329 | cg27179424 | *HDLBP* | -1630 | 276 | 5.23e-08 |  | NA | NA | NA |
| *TOMM40* | 8 | 145003862 | cg04757492 | *PLEC* | -1850 | 318 | 8.29e-08 |  | 710.00 | 349.00 | 0.04 |
| *TOMM40* | 8 | 145813981 | cg11488020 | *ARHGAP39* | -2110 | 361 | 7.15e-08 |  | 829.00 | 349.00 | 0.02 |
| *TOMM40* | 9 | 139835353 | cg14024965 | *FBXW5* | -1890 | 306 | 1.52e-08 |  | 1020.00 | 390.00 | 0.01 |
| *TOMM40* | 11 | 1431643 | cg13767324 | *BRSK2* | -2180 | 378 | 9.4e-08 |  | NA | NA | NA |
| *TOMM40* | 11 | 1581959 | cg17500385 | *MOB2_DUSP8* | -1760 | 297 | 4.97e-08 |  | NA | NA | NA |
| *TOMM40* | 11 | 3059371 | cg16730141 | *CARS1* | -2260 | 373 | 2.56e-08 |  | NA | NA | NA |
| *TOMM40* | 16 | 833382 | cg08469326 | *MSLNL_RPUSD1* | -1540 | 250 | 1.65e-08 |  | NA | NA | NA |
| *TOMM40* | 18 | 3594197 | cg03760316 | *DLGAP1* | 7360 | 1210 | 2.25e-08 |  | -3780.00 | 913.00 | 4.15E-05 |
| *TOMM40* | 19 | 2120959 | cg24438277 | *AP3D1* | -2310 | 397 | 7.69e-08 |  | 797.00 | 391.00 | 0.04 |
| *TOMM40* | 19 | 8464850 | cg11817453 | *RAB11B* | -1820 | 313 | 8.05e-08 |  | 607.00 | 339.00 | 0.07 |
| *TOMM40* | 20 | 62052259 | cg13379325 | *KCNQ2* | -1710 | 283 | 2.88e-08 |  | 562.00 | 320.00 | 0.08 |
| *APOE* | 3 | 49756571 | cg15078681 | *RNF123_AMIGO3* | -46300 | 7290 | 7.09e-09 |  | -7220.00 | 8360.00 | 0.39 |
| *APOE* | 11 | 1430600 | cg11285322 | *BRSK2* | 40300 | 6750 | 3.9e-08 |  | NA | NA | NA |
| *APOE* | 14 | 104178543 | cg11718757 | *XRCC3_KLC1* | -53700 | 8960 | 3.71e-08 |  | NA | NA | NA |
| *APOE* | 19 | 48000364 | cg24009074 | *NAPA* | 41700 | 6750 | 1.55e-08 |  | 4600.00 | 11200.00 | 0.68 |
| *APOE* | 20 | 31127002 | cg08006263 | *NOL4L* | 53700 | 8910 | 3.18e-08 |  | NA | NA | NA |

*BETA, STDERR, and *P* represent the regression coefficient and its corresponding standard error, and *P* value of the generalized linear mixed regression model where the DNA methylation level of the CpG site was exposure variable and the mRNA expression level of the targeted gene was the outcome variable with the adjustment of the fixed covariates of age at death, sex, technical variables of batch and methylation array position, and the random covariate of methylation chip.
